# Moving together – Benefits of a 12-week online dance training intervention on static and dynamic postural stability and gait speed in older adults – a pre-post study

**DOI:** 10.1101/2023.11.01.23297899

**Authors:** Rasmus Kopp Hansen, Elizabeth Jochum, Ditte Egholm, Morten Villumsen, Rogerio Pessoto Hirata

## Abstract

**Background:** Physical inactivity negatively affects gait performance and postural stability in older adults, which results in a higher risk of fall accidents. Previous research has shown that in-person dance training improves various aspects of balance and lower extremity function, however, the potential benefits of dance training delivered online on variables used for fall risk stratification in older adults have yet to be studied. We aimed to explore the benefits of a 12-week online dance training intervention on static and dynamic postural stability and gait speed in older adults.

**Methods:** Forty-five older adults were included. The 12-week dance training intervention consisted of two weekly 60-min classes in improvisation and salsa delivered online through Zoom video calls. Static and dynamic postural stability was assessed using the center of pressure (CoP) area and velocity (force platform), and the Mini Balance Evaluation Systems Test (Mini-BESTest), respectively. 10-m gait speed was measured using photo gates. Before and after comparisons were performed using paired sample t-tests.

**Results:** Thirty-two older adults completed the study. There were no significant changes for static postural stability assessed by CoP area or velocity (P ≥ 0.218). The Mini-BESTest total score was significantly improved at post-intervention (23.88 ± 3.01) compared to baseline (22.56 ± 1.41, P = 0.007). 10-m gait speed was significantly faster at post-intervention (1.68 ± 0.25 m/s) compared to baseline (1.57 ± 0.22 m/s, P < 0.001).

**Discussion:** These results indicate that online dance training has limited effect on static postural stability but may be beneficial for gait speed and in particular dynamic postural control among older adults. While the absolute increase in gait speed observed in the present study suggests limited clinical relevance, the improvement in Mini-BESTest score was above the minimal clinically important difference, which suggests a clinically relevant enhancement of dynamic postural control.

## Introduction

Physical inactivity negatively affects gait performance and postural stability in older adults, which results in a higher risk of fall accidents. Medical costs related to falls in older adults were estimated to be approximately $50 billion per year in the USA alone (1). This is a concerning public health problem, as around 30% of the older adult population is expected to experience an accidental fall within a one-year time period (2). Common outcomes from fall accidents in older adults are: head injuries (3), fear of falling (4), loss of physical independence (5), increased healthcare utilization (6), and mortality (7), among others.

Physical training has been shown to improve risk factors and prevent fall accidents among older adults by focusing on improving gait and balance stability deficits (8). In the current world guidelines for fall prevention, dynamic gait evaluations are used for stratification of older adults in relation to fall risk (9). The Mini Balance Evaluation Systems Test (Mini-BESTest) has been extensively used in clinical settings to evaluate anticipatory and reactive postural responses and gait dynamics (10) due to its high construct validity and ability to discriminate between groups with respect to fall risk in older adults (11). Furthermore, low gait speed has previously been correlated with increased risk for accidental fall in community-dwelling older adults (12), a risk that is specifically amplified when gait speed is lower than 1 m/s (13). Jointly, these studies indicate the importance of physically active aging in preventing fall accidents.

According to systematic reviews, dance training is a valuable health promotion intervention for supporting healthy aging and addressing risk factors with declining health (14), and for mitigating fall risk (15). Specifically, dancing is an enjoyable physical activity that can be tailored to different cultural and aging backgrounds, and the physical needs of older adults; can improve physical health outcomes; and has the potential to enhance emotional, psychological, and social well-being. While Meron et al. (16) reported that folk or ballroom dancing did not prevent falls or associated physical and cognitive fall-related risk factors, other studies using in-person dance training have reported improved medial-lateral balance (17), reduced dependency of visual input for balance maintenance (18), reduced attention demands during postural tasks (17), and improved lower extremity function in older adults after as little as one weekly dance class for 12 weeks (19). In addition, a feasibility study (20) recently demonstrated beneficial impact of an 8-week contemporary dance training program on the modification of both physical and psychological risk factors for falling in community-dwelling older adults. Taken together, although accumulating evidence supports the benefits of in-person dance training on healthy aging, including the risk of falls, virtually no studies have explored the potential benefits of dance training delivered digitally (online) (21). Such online-based dance training may be particularly relevant in times of restricted access to physical activity, such as during the recent COVID-19 pandemic. Moreover, while home-based and digital training have previously proven to be effective for combating physical inactivity among this population (22), less is known about whether this type of training may improve variables used for fall risk stratification in older adults, such as gait speed and postural stability.

Therefore, the aim of the study was to explore the benefits of a 12-week online dance training intervention the *Moving Together* (MT) program, on static and dynamic postural stability and gait speed. Static postural stability is here defined as the ability to hold an upright position as still as possible over a stable base of support, while dynamic postural stability is the ability to maintain balance, i.e., keeping the body’s center of mass vertical projection within the base of support, when changes in the base of support occur (23). Throughout the evaluation of the intervention’s impact on these specific physical aspects, the study also seeks to contribute valuable insights into the literature on the potential benefits of online dance training in the functional performance of older adults. The hypothesis was that static postural stability, dynamic postural stability, and gait speed would improve after 12 weeks of the online MT intervention.

## Materials and Methods

### Participants

Forty-five older adults were included in this uncontrolled pre-post study (44 females; 74 ± 5.3 years; 69.7 ± 13.0 kg; 1.62 ± 0.58 m). The participants were recruited through advertisements posted at activity centers for older adults in Aalborg Municipality, through information meetings, by advertisement in the press, and through flyers handled out in public spaces. To ensure inclusivity, only two inclusion criteria were used: I) being 65 yrs or older, and II) being able to speak and understand Danish. Due to the wide inclusion criteria, there was a broad age range for participants (65 to 87 yrs) as well as physical capacity of the participants. However, all were able to stand and walk independently. The study was approved by the North Denmark Region Committee on Health Research Ethics (N-20220045) and conducted in accordance with the Declaration of Helsinki.

### Dance training intervention

The study builds upon a previous randomized controlled trial from our group (24) demonstrating that regular fitness circuit training and a combination of in-person and online dance training significantly reduced the number of fall accidents in older adults (25). In this separate study, we sought to extend this work by focusing on implementation of online dance training in practice (in the municipality). Specifically, the MT consisted of two weekly dance classes lasting 60 min each, consisting of improvisation (contemporary dance) (Day 1) and salsa (Day 2) performed for 12 weeks. The movement language of Laban’s choreological practice (26) was used to inspire and create variations of movement during the contemporary dance classes, and the classes progressed in intensity. Each class started and concluded with a focus on breathing exercises. The salsa classes were built upon techniques from Cuban salsa; participants had a progressive introduction to the technique and the expressive aesthetics of the dance style. Over time, the class included dancing with faster pace. The classes were delivered by experienced professional dance instructors online through video calls using the Zoom meeting application (Zoom Video Communications, Inc., San Jose, CA, USA). For flexibility purposes, subjects could choose to attend the online classes either from home, or at one of four activity centers in the municipality. Each online class was recorded and uploaded to a designated YouTube channel connected to the project after each class._^1^_ The YouTube channel resource was made available for all participants and the public, where participants could review and replay the classes if they were unable to attend on the day. Participants were instructed to dance at least once per week but were encouraged to attend twice weekly. To promote engagement and social connection between the participants, a 60-min in-person salsa dance class was conducted once every month with physical presence by both the instructor and the participants (up to three sessions in total per subject).

### Measurements

All measurements were performed immediately prior to (baseline) and following 12 weeks of dance training and were carried out at local activity centers in Aalborg Municipality. The same experienced investigator was present at all tests and either performed the testing or provided supervised assistance to two student helpers.

#### Static Postural Stability

Static postural stability during a quiet standing task was assessed for 30 seconds both with and without a dual-task. A high-resolution three dimensional, four-channel, force platform (PLUX Wireless Biosignals S.A., Lisboa, Portugal) sampled at 1000Hz was placed 1.5 m from a wall. A circle (d = 5 cm) was placed on the wall at approximately eye height straight out of the participants. At baseline, participants were instructed to stand on the force platform in a self-determined, comfortable position with the distance between the feet measured with a non-elastic measuring tape. The distance between the feet was then replicated at the post-test. Three single- and three dual-task recordings were performed, and participants were instructed not to move during the 30-s recording period. Measurements were calibrated prior to each participant, by recording a 10-s period with zero load on the force platform. The center of pressure (CoP) data from the force platform was extracted from the horizontal forces and filtered digitally with a low-pass filter and a 20Hz cut-off frequency. The static postural stability was then quantified from the CoP data via the 95% prediction ellipse CoP area and resultant mean CoP velocity (27). A mathematic task was used as supra-postural task during the dual-task conditions, when participants were instructed to count backwards of ‘minus 7’, starting from a random number between 0 and 500 (e.g., 40, 33, 26 etc.). The starting number for the three dual-tasks remained constant between visits. The mean of the three single-tasks and the three dual-tasks for both variables were used for analyses.

#### 10-m gait speed

Gait speed was determined using photo gates (Brower Timing Systems, Draper, UT, USA) placed in a corridor 10 meters apart. Participants were instructed to walk as fast as possible without running (“Please walk as quickly as you can without running to the end of the corridor”). Participants started walking two meters before the start line so that gait speed did not include the acceleration time, with an additional 2-m walk-out after the participants crossed the finish line. A total of six walks were performed (6 × 10 meters), with a self-determined break in between walks. For the last three 10-m walks, participants were instructed to perform the same mathematical dual-task as for the assessment of postural control, yet with other starting numbers. For these dual-tasks, participants were instructed to walk as fast as possible without running, while counting as correctly as possible. The mean of the three single-tasks and the three dual-tasks, respectively, were used for analyses. For comparison with previous studies, the time to complete the 10-m walk was converted to speed (m/s).

#### Dynamic postural stability (Mini-BESTest)

Dynamic postural stability was determined using The Mini-BESTest (10). The Mini-BESTest consists of 14 items related to domains of anticipatory transitions; reactive postural control; sensory orientation; and dynamic balance during gait. The procedures for administration and scoring of the Mini-BESTest were standardized as described in (28). For each item, a score from 0 (severe) to 2 (normal) was provided. Individual item scores were then summated and reported as the Mini-BESTest total score (out of the maximum attainable 28 points) (28).

### Statistics

Statistical analyses were performed using SPSS software (version 27; IBM, Armonk, New York) with data reported as mean ± SD. Statistical significance was set *a priori* at P < 0.05. Before and after comparisons on outcomes were performed using student paired t tests (two-tailed), with Bonferonni adjustment for multiple comparisons (n = 2; gait speed with and without dual-task) by dividing 0.05 with the number of comparisons such that significance was accepted at P < 0.025 for these two outcomes. Pearson product-moment correlations (or Spearman’s rank-order correlation for non-normally distributed change scores) were calculated between age and change scores (Δ = post-intervention - baseline) for outcomes to explore potential associations between age and responses to the intervention. We also calculated standardized effect sizes (Cohen d) to determine the magnitude of change from baseline to post-intervention, with the following thresholds used for interpretation: small (>0.2), moderate (>0.5), and large (>0.8).

## Results

Of the 45 participants included at baseline, 13 dropped out, so that n = 32 in total were included for analyses. All 32 participants were included in analyses irrespective of where and how they attended. Reasons for dropout included non-study related health issues (n = 5), lack of motivation (n = 4), COVID-19 infection (n = 1), finding it too physically demanding to complete the dance sessions (n = 2), and finding it uncomfortable being the only male (n = 1).

### Static Postural Stability

Results for static postural stability are shown in Table 1. Due to corrupted data for three participants, analyses are based on n = 29. There were no significant changes from baseline to post-intervention for either single-or dual-task CoP area or velocity (P ≥ 0.218). Due to non-normality of change scores, Spearman’s rank order correlations were run to assess relationships between age and Δ in single and dual-task CoP area and velocity. There were statistically significant correlations between age and Δ in CoP area during single (r_s_ = 0.386, P = 0.038) and dual-task (r_s_ = 0.425, P = 0.022), suggesting that older age was associated with a greater increase in CoP area over the 12 weeks. Similarly, there was a statistically significant correlation between age and Δ in dual-task CoP velocity (r_s_ = 0.425, P = 0.021), with no correlation between age and Δ in single-task CoP velocity (r_s_ = 0.191, P = 0.321). The mathematical performance during the dual-task is presented in Table 2. There were no significant changes in the mathematical performance during the dual-task, either when expressed as the total number of counts, or as the relative or absolute number of correct counting’s (P ≥ 0.28).

**Table 1.** Static postural stability assessed via area and velocity of center of pressure (CoP) during 30s of quiet standing with and without a dual mathematical load at baseline and after 12 weeks of online dance training (n = 29*)

|  | Baseline | Post-intervention | P-value | Effect size (d) |
| --- | --- | --- | --- | --- |
| **CoP Area (cm<sup>2</sup>)** |  |  |  |  |
| Single task | 2.50 ± 2.43 | 2.08 ± 1.33 | 0.351 | 0.176 |
| Dual task | 3.83 ± 3.26 | 3.47 ± 3.43 | 0.430 | 0.149 |
| **CoP Velocity (cm/s)** |  |  |  |  |
| Single task | 1.29 ± 0.42 | 1.28 ± 0.50 | 0.889 | 0.026 |
| Dual task | 1.70 ± 0.69 | 1.90 ± 1.06 | 0.218 | 0.234 |

\* Due to corrupted data for three participants, n = 29.

**Table 2.**
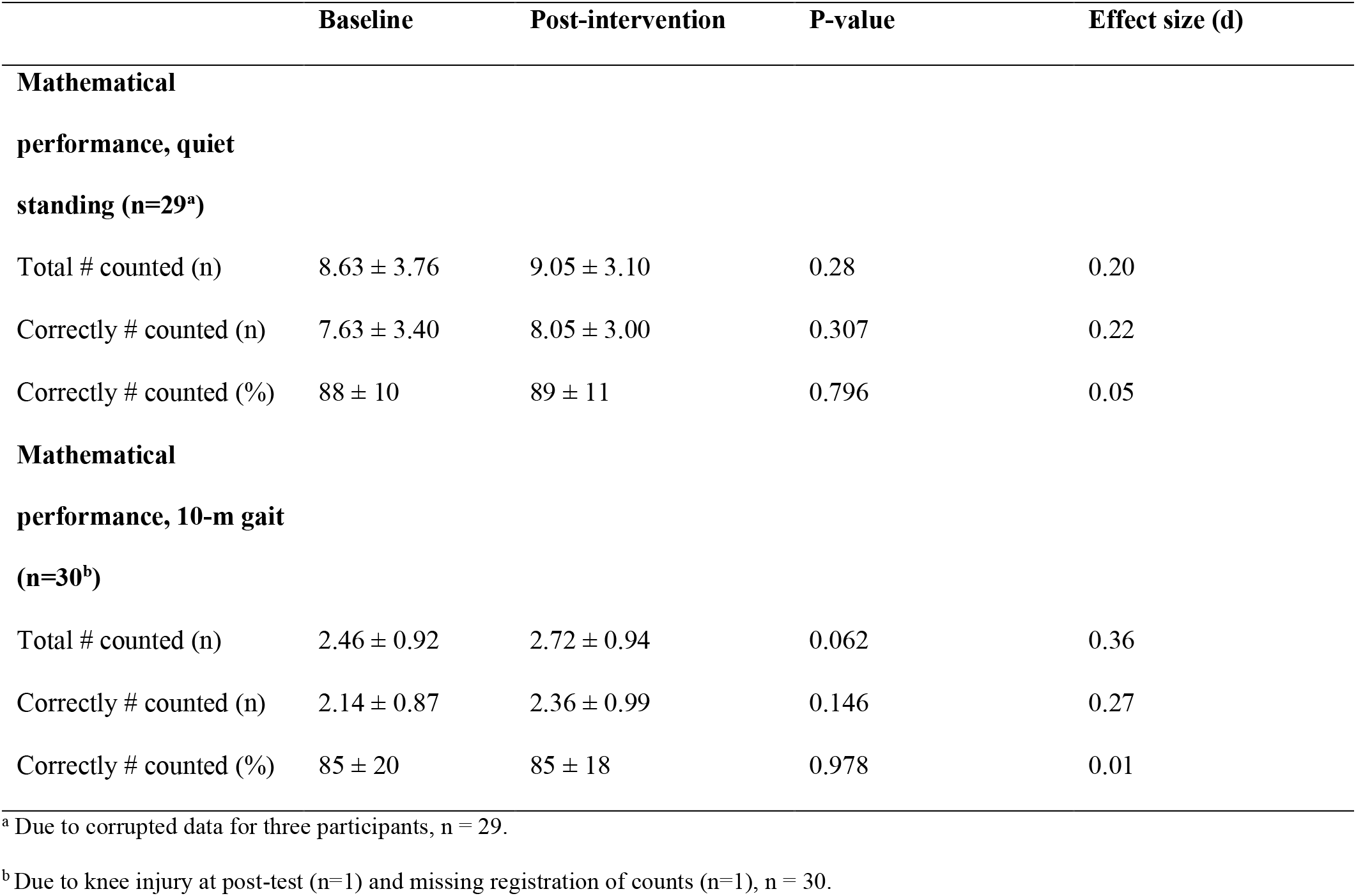
Performance during the dual-tasks consisting of respectively 30s of quiet standing and 10-m gait with a dual mathematical load (counting backwards of minus 7) at baseline and after 12 weeks of online dance training.

### 10-m gait speed

One participant was unable to complete the walking test at post-intervention, so for 10-m gait speed analyses is based on n = 31. 10-m gait speed was significantly faster after the 12 weeks of dance training (1.68 ± 0.25 m/s) compared to baseline (1.57 ± 0.22 m/s, P < 0.001, d = 0.737, Figure 1A). After Bonferroni adjustment, there was no significant change in 10-m gait speed with dual-task from baseline (1.41 ± 0.21 m/s) to post-intervention (1.48 ± 0.25 m/s, P = 0.045, d = 0.376, Figure 1B). There were no significant correlations between age and Δ in 10-m gait speed (r = -0.072, P = 0.699) or age and Δ in 10-m gait speed with dual-task (r = -0.99, P = 0.595). The mathematical performance during the dual-task is presented in Table 3.

**Figure 1.**
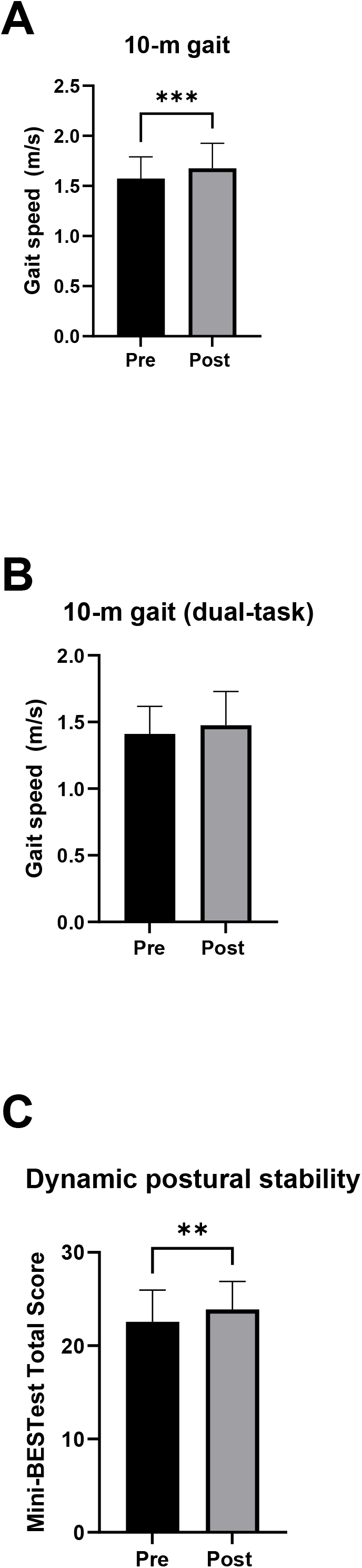
Mean gait speed and dynamic postural stability at baseline (Pre, black bars) and following 12 weeks of dance training (Post, grey bars). (A) 10-m gait speed. (B) 10-m gait speed with a mathematical dual-task. (C) Mini-BESTest Total Score. Mini-BESTest Total Score was calculated as the sum of the 14 test items. Note that for gait speed, n = 31. Error bars represent SD. Significant difference between baseline and post-intervention at **P <0.01 and ***P <0.001.

### Dynamic Postural Stability

Dynamic postural stability evaluated by the Mini-BESTest total score was significantly improved at post-intervention (23.88 ± 3.01) compared to baseline (22.56 ± 1.41, P = 0.007, d = 0.52, Figure 1C). There was no significant correlation between age and Δ in Mini-BESTest total score (r = 0.085, P = 0.642).

## Discussion

When comparing the results after 12 weeks of the MT with baseline values, our initial hypotheses were only partially confirmed: I) it is not clear that static postural stability improved after the online dancing intervention, as both the CoP area and velocity variables were not significantly different between before and after intervention, II) gait speed was only faster during the conditions without dual-task, however III) dynamic postural control, determined via the Mini-BESTest, was enhanced following the intervention.

### Static Postural Stability

Assuming that static postural stability during a quiet standing task is achieved via controlling the body posture as if it behaves as an inverted pendulum swinging around the ankle joints (29), the optimal neuromuscular strategy to mitigate the effect of possible external perturbation is to increase the stiffness around the ankles by co-contracting the antagonistic and agonistic muscles around this joint (30). This strategy usually results in smaller and quicker displacements of the body after a perturbation, which, in laboratory settings, are usually represented by smaller areas and increased velocities of the CoP (31). Adding a supra-postural task, older adults would exhibit a decreased (I) static postural stability (32), or (II) performance of the supra-postural task when the postural task is highly demanding (33). In older adults, the capacity to carry out concurrent postural tasks is paramount for maintaining balance in daily life activities, since its deficit may increase the risk for accidental falls in this population (34). This capacity may also involve the prioritization of one of the tasks over the other, which in postural control studies is often referred to as the “postural first” principle, when the focus on maintaining the static posture stability overcomes the performance of the supra-postural task (33). Regardless of the addition or not of a supra-postural task, the present results did not show any clear change in CoP area or velocity nor in the performance of the dualtask intervention before/after. This could indicate: (I) a plateau effect, where the older adults that participated in this study already presented good static postural stability prior to the intervention, (II) a sensitivity problem, where the force platform measurement used here where not able to detect the possible variations due to training, or (iii) ineffective intervention for static postural stability since the dance training focused primarily on dynamic movements which relate to dynamic postural stability. As exploratory analyses, we also computed correlations between age and changes in postural control and gait speed to explore whether changes in these outcomes would be dependent on age. For these analyses, we found significant correlations between age and Δ in CoP velocity (single-task only) and for age and Δ in CoP area and velocity (both single and dual-task), thus largely confirming previous research demonstrating greater CoP parameters at older age during single-(35) and dual-task (36) platform assessments.

### Dynamic Postural Stability

The ability to control the body position in space so its center of mass is within the base of support when the body suffers perturbation (dynamic balance) relies upon the available sensory information (usually vision, proprioceptive, and vestibular) to generate accurate motor responses to counter-react such perturbation and realign the vertical projection of the bodies’ center of mass within the new base of support. Such a scenario is often encountered when performing daily life activities such as walking, running, dancing, carrying objects, using transportation, doing house chores, etc. The level of performance of such activities is usually associated with accidental falls in older adults, where lower independence in performing these activities is associated with a higher risk of fall accidents in older adults (37). After 12 weeks of the online dance training intervention, the Mini-BESTest score was significantly greater compared to baseline values, with a moderate effect size.

These results indicate that the MT intervention was beneficial for improving the older adults’ dynamic postural stability. It is noteworthy that the baseline values for Mini-BESTest (average of 22.5) were below the cutoff value of 23 points for the average age (74 years) reported in the literature (38), which indicates that on average, participants were in a higher risk for fall accidents when enrolled in the study compared to their peers. After the intervention, the average score for this test increased to approximately 24 (with an average increase of 1.5 points), which is not only above the cutoff values for fall risk in that age group, but also similar in size to the minimal clinically important difference (1-2 points on the Mini-BESTest scale) when evaluating special groups such as individuals after total knee replacement (39). However, it is important to highlight that the number of fall accidents where not monitored in this study, so the real effect on fall accidents from this intervention is yet to be investigated.

### Gait Speed

Gait is also an effective way to quantify dynamic postural stability in older adults (40). In fact, increasing gait speed after training has been correlated with lower fall risks in older adults. In average, the gait speed increased 0.1 m/s after the present intervention, which even though representing a moderate change (d = 0.737) might not be clinically relevant. The lack of relevance of this result might be related to a ceiling effect. The maximum gait speed in older populations is expected to range from 1.7 m/s (females) to 2.1 m/s (males) (41), while the average gait speed during the baseline test was 1.5 m/s (all females). It is noteworthy that according to the latest recommendations (9), gait speed should be used for fall risk stratification among older adults.

Indeed, gait speeds below 1 m/s are related to a higher risk of accidental falls in older adults (12), indicating that for our participants, gait speed would not be an efficient measure for fall risk stratification. Nevertheless, after the online dance intervention, our participants reached values close to the maximum (1.6 m/s), indicating that the intervention may have provided enough stimuli for motor improvement even though the participants already exhibited high performance values at baseline. Further studies should investigate whether an online training approach is also effective for less capable participants.

### Methodological considerations

Despite the high degree of external validity in this study as the intervention was implemented in practice, in a municipal setting, the study is limited, however, by not including a control group for comparison. Thus, the results from the current study should be interpreted with some caution.

Another consideration is that our results might not be generalizable to older males, as only one man was included in the study, and he dropped out due to not feeling comfortable with being outnumbered. Such overwhelming female bias is consistent with the existing literature (14,42) and should be addressed in future studies. Finally, this study is limited by the lack of control of participant attendance. Due to our focus on inclusivity and implementation, participants were allowed to play and replay the recorded dance classes from YouTube on their own, and hence we were unable to accurately record attendance. This may have resulted in some participants attending only once weekly, while others may have attended more than twice weekly. While we acknowledge this limitation, it may approximate the more real-life context of our study.

## Conclusion

In summary, while the MT online dance training intervention had a limited impact on static postural stability and gait speed, our data indicate important benefits of the intervention for dynamic postural control among older adults. Further research, including studies with stricter control of study variables, is needed to investigate the efficacy of online dance training on the number of fall accidents and to explore its effectiveness with different participant groups.

## Data Availability

All data produced in the present study are available upon reasonable request to the authors

## Acknowledgments

The authors would like to thank Monica Tøsti Christensen and Mads Præstegaard Sørensen for their help with data collection; Frederikke Spedsbjerg for maintenance of the YouTube channel; The Association of Integrated Dance in Denmark for their input to the dance classes; and all the participants for their involvement in the study.

## Footnotes

1 https://www.youtube.com/@movingtogether8461

